# A Wastewater-based Epidemiology tool for COVID-19 Surveillance in Portugal

**DOI:** 10.1101/2021.07.21.21260905

**Authors:** Sílvia Monteiro, Daniela Rente, Mónica V. Cunha, Manuel Carmo Gomes, Tiago A. Marques, Artur B. Lourenço, Eugénia Cardoso, Pedro Álvaro, Marco Silva, Norberta Coelho, João Vilaça, Fátima Meireles, Nuno Brôco, Marta Carvalho, Ricardo Santos

## Abstract

Shedding of severe acute respiratory syndrome coronavirus 2 (SARS-CoV-2) RNA in the feces and urine of infected patients and subsequent presence in wastewater has produced interest on the use of this matrix for sentinel surveillance at a community level and as a complementary approach to syndromic surveillance. With this work, we set the foundations for wastewater-based epidemiology (WBE) in Portugal by monitoring the trends of SARS-CoV-2 RNA circulation in the community, on a nationwide perspective during different epidemiological phases of the pandemic. The Charité assays (E_Sarbecco, RdRP, and N_Sarbecco) were applied to monitor, over 32-weeks (April to December 2020), the dynamics of SARS-CoV-2 RNA at the inlet of five wastewater treatment plants (WWTP), which together serve more than two million people in Portugal. Raw wastewater from three COVID-19 reference hospitals was also analyzed during this period. In total, more than 600 samples were tested. Sampling started late April 2020, during lockdown, and, for the first weeks, detection of SARS-CoV-2 RNA was sporadic, with concentrations varying from 10^3^ to 10^5^ genome copies per liter (GC/L). Prevalence of SARS-CoV-2 RNA increased steeply by the end of May into late June, mainly in Lisboa e Vale do Tejo region (LVT), during the reopening phase. After the summer, with the reopening of schools in mid-September and return to partial face-to-face work, a pronounced increase of SARS-CoV-2 RNA in wastewater was detected. In the LVT area, SARS-CoV-2 RNA load agreed with reported trends in hotspots of infection. Synchrony between trends of SARS-CoV-2 RNA in raw wastewater and daily new COVID-19 cases highlights the value of WBE as a surveillance tool for this virus, particularly after the phasing out of the epidemiological curve and when hotspots of disease re-emerge in the population which might be difficult to spot based solely on syndromic surveillance and contact tracing.

## 1. Introduction

Climate change, deforestation and population growth led to an increase in contact between humans and wildlife, which may cause interspecies transmission of infectious agents. Such conditions possibly resulted in the occurrence of previous outbreaks including the severe acute respiratory syndrome (SARS; 2002-2004) and the Middle East respiratory syndrome (MERS; 2012-present) outbreaks, all caused by coronavirus (CoV; SARS-CoV and MERS-CoV, respectively). Several authors that have addressed the environmental circulation of viruses had already highlighted the possible occurrence of a new pandemic caused by coronavirus (Wigginton and Ellenberg, 2015; Santos and Monteiro, 2013).

Coronavirus disease 2019 (COVID-19) is caused by the severe acute respiratory syndrome coronavirus 2 (SARS-CoV-2), an enveloped, single-stranded RNA virus with high infection rate. The first clinical cases in Portugal were reported on March 2, 2020, entering the exponential phase on March 14, 2020 (RTP, 2020). The Portuguese government closed schools on March 16, 2020, and declared the emergency state on March 19, 2020, with the country entering the first national lockdown until May 2, 2020. Reopening occurred in three stages throughout the month of May, with full reopening in June 2020 except for schools that remained closed throughout the month. In September, schools reopened, and partial face-to-face work returned, leading to a steep increase in the number of cases (DGS, 2020). As of December 2, 2020, 73,876 COVID-19 cases had been reported in Portugal, with 4,724 deaths and 229,018 recovered patients (DGS, 2020).

Although COVID-19 clinical tests have been developed in record time, the disease spread, and community infection burden often outpaced the capacity for clinical testing. In addition, syndromic surveillance strongly depends on individual reporting and seriousness of clinical symptoms, and how this coincides with diseases known to circulate in the community (Mandi *et al*., 2020). Rapid approaches to determine the extent of virus spread in the population, ideally in near real-time, are thus needed to slow down transmission.

Wastewater-based epidemiology (WBE) has been applied since 2005 to trace pharmaceutical and illicit drug use in the community (Zuccato *et al*, 2005; Reddy, 2010; Singer *et al*., 2013; Choi *et al*., 2018). The usefulness and potential of wastewater as a surveillance system for pathogens has already been shown, namely under the global polio eradication initiative, the most successful example of environmental surveillance to date (Hovi *et al*., 2012; WHO, 2015; Koopmans *et al*., 2017).

Several advantages are associated with WBE; firstly, testing wastewater means testing thousands of potentially infected individuals at the same time, with the capacity to identify hotspots of infection prior to symptomatic surveillance. Secondly, WBE can highlight trends in virus shedding over time from symptomatic but also from asymptomatic, pre-symptomatic and post-symptomatic individuals.

SARS-CoV-2 although transmitted mainly via respiratory droplets (Meselson, 2020). has been detected in feces and urine of infected patients, regardless of disease severity or development of gastrointestinal illness (He *et al*., 2020; Pan *et al*., 2020; Wölfel *et al*., 2020; Young *et al*., 2020). Although there is little indication that virus shed in the stools of infected patients, and therefore circulating in wastewater, are infectious (Wölfel *et al*., 2020; Zang *et al*., 2020), the presence of SARS-CoV-2 RNA in raw wastewater provides valuable information regarding the emergence, prevalence, epidemiology and decrease of SARS-CoV-2 presence in the community, helping the early identification of hotspots of infection.

To date, several authors reported the occurrence of SARS-CoV-2 RNA in wastewater samples (Ahmed *et al*., 2020; Medema *et al*., 2020; Randazzo *et al*., 2020; Sherchan *et al*., 2020) demonstrating the usefulness of WBE for SARS-CoV-2. Several iterations of the application of WBE for SARS-CoV-2 are currently implemented in many countries, such as the Netherlands, Scotland and Spain among others.

In this study, we report for the first time the results of SARS-CoV-2 RNA monitoring in raw wastewater in Portugal, in a study covering about 20% of the Portuguese population, corresponding to over two million people, over a 32-weeks period. More than 600 samples were collected from five wastewater treatment plants (WWTP) and three COVID-19 hospitals in two regions of the country, a north cluster (four municipalities) and a south cluster in Lisboa e Vale do Tejo (LVT) (six municipalities) To the best of our knowledge, this is the first study jointly evaluating the presence of SARS-CoV-2 RNA in raw wastewater from WWTP and COVID-19 hospitals while encompassing distinct epidemiological phases of the pandemic as well.

## 2. Materials and Methods

### 2.1. Clinical surveillance data

Clinical surveillance data were obtained from the Reports from the Portuguese Health Authority (DGS, 2020). Data from clinical surveillance for each municipality were presented daily in the reports from the Health Authority, being provided on a weekly basis after July 2020.

### 2.2. Sampling sites and sample collection

Raw wastewater samples (*n* = 404) were collected between April 27, 2020, and December 2, 2020, from five WWTP located in the North (Gaia Litoral (GA) and Serzedelo II (SE)) and in LVT (Alcântara (AL), Beirolas (BE), and Guia (GU)) (Fig. S1) of Portugal. Further information about these WWTP catchments is provided in Table S1. Sampling took place for 102 days, covering 220 of calendar days in total. Raw wastewater from three reference COVID-19 hospitals (Hospital Curry Cabral (HCC), Lisbon; Hospital Sra. Oliveira (HSO), Guimarães (North); and Hospital Santos Silva (HSS), Vila Gaia (North); *n* = 204), in the catchment area of the WWTP, was also sampled.

Twenty-four-hour composite samples were collected using automated samplers (ISCO, US), except for HSO and HSS, where due to logistical problems only grab samples were taken. Samples were transported refrigerated to the laboratory, within 8 h of collection and processed immediately upon arrival to the laboratory.

### 2.3. Processing of raw wastewater

Upon arrival to the laboratory, 1-L of raw wastewater from WWTP and COVID-19 hospitals was concentrated using hollow-fiber filters Inuvai R180 (Inuvai, a division of Fresenius Medical Care, Germany). Samples were eluted in 300 mL of 1X PBS containing 0.01% NaPP and 0.01 Tween 80/0.001% antifoam and precipitated overnight with 20% polyethylene-glycol (PEG) 8000. Samples were then centrifuged at 10000 xg for 30 min and resuspended in 5 mL 1X PBS, pH 7.4 (Blanco *et al*., 2019). Samples were kept at (−80 ± 10) ºC until further processing.

### 2.4. Viral extraction, detection, and quantification

Viral RNA was extracted from 220 µL concentrated samples using the QIAamp FAST DNA Stool Mini kit (QIAGEN, Germany), according to the manufacturer’s instructions. The RNA was recovered in a final volume of 100 µL.

Primers and probes used in this study are presented in Table 1. The recovery efficiency for RNA extraction was performed using murine norovirus (MNV), which was added to the concentrates as an extraction control. MNV RNA was detected and quantified using the assay described by Baert *et al*., 2008. SARS-CoV-2 RNA was detected using the Charité assays: the E_Sarbecco, targeting the envelope protein gene, the RdRp that targets the RNA-dependent RNA polymerase gene and the N_Sarbecco, which targets the nucleoprotein (Corman *et al*., 2020).

**Table 1.**
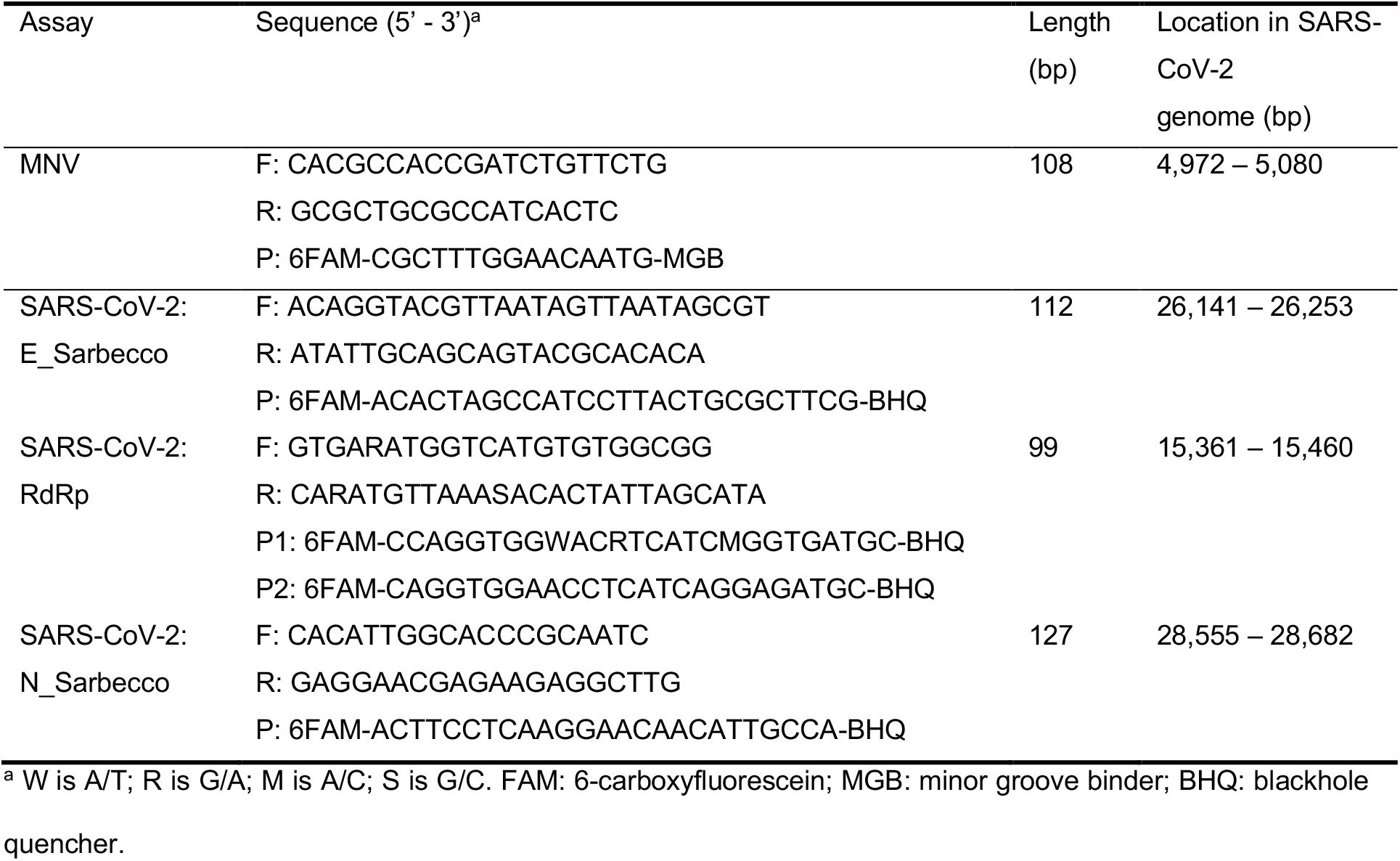
Primers and probes used in this study

One-step RT-qPCR assays (AgPath-ID™ One-Step RT-PCR, Thermo Scientific, USA) was used for the quantitative detection of SARS-CoV-2 and MNV. For the specific detection and quantification of viral RNA, 5 µL of 4-fold and 10-fold dilutions of each viral RNA extract were also assayed in parallel with crude extracts; dilutions were meant to overcome amplification inhibition due to the complex nature of the samples. The final volume of reaction mixture was 25 µL, composed of 800 nM of each primer, 200 nM of probe and 5 µL of extracted RNA. RT-qPCR reactions were carried out at 45 ºC for 10 min, 95 ºC for 10 min, followed by 45 cycles of amplification at 95 ºC for 15 s and 58 ºC for 45 s for SARS-CoV-2 and 60 ºC for 45 s for MNV. RT-qPCR was performed on an Applied Biosystems 7300 Real-Time PCR System (Applied Biosystems, US). Reactions were considered positive only if the cycle threshold was below 40 cycles (Medema *et al*., 2020; F. Wu *et al*, 2020). Quantification of E_Sarbecco and RdRp assays was performed through calibration curves using 10-fold dilutions of nCoV-ALL-Control plasmid (Eurofins Genomics, Germany), ranging from 1.94 to 1.94 × 10^6^ and 1.00 to 1.00 × 10^6^ GC per reaction respectively. Quantification of N_Sarbeco assay was performed using 2-fold and 10-fold dilutions (ranging between 2.00 to 2.00 × 10^4^ GC per reaction) of the Amplirun SARS-CoV-2 RNA control (Vircell, Spain). Negative controls (extraction and RT-qPCR assay) were also performed using DNase/RNase free distilled water, following the same conditions as the samples. The extraction efficiency using MNV as proxy averaged 70% (±19%).

### 2.5. SARS-CoV-2 RNA load estimates standardized to population

Standardization of SARS-CoV-2 RNA concentration to population and WWTP for each sampling date was performed in accordance with Eq. 1 (Gonzalez *et al*., 2020). For this calculation only the results from E_Sarbecco assay were used since it was the most sensitive assay.

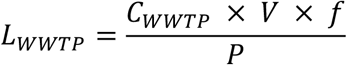

where:

*L*_*WWTP*_ is SARS-CoV-2 RNA load in the WWTP standardized to the population (GC per person in the catchment)

*C*_*WWTP*_ is the SARS-CoV-2 RNA concentration in samples yielded by the E_Sarbecco assay (GC/L)

*V* is the average daily flow of wastewater in the WWTP during the sampling day (m^3^/day)

*f* is the conversion factor between L and m^3^

*P* is the estimated population within the WWTP catchment.

### 2.6. Data analysis

All data analysis was done with SPSS version 26 (IBM Corporation, US). For statistical analysis, all RT-_q_PCR below the limit of detection (LOD) were substituted by the LOD with subsequent log_10_ transformation. The LOD was 3.99, 5.52 and 5.74 GC per reaction for E_Sarbecco, RdRp and N_Sarbecco assays, respectively. Kruskal-Wallis test (KW statistics) was conducted to compare differences in the total number of SARS-CoV-2 RNA detection for each assay, and pairwise comparison was performed with Dunn’s test. Mann-Whitney test was used to determine the impact of sampling type (composite *versus* grab samples collected at hospitals). Spearman rank order correlation was used for calculation of correlation coefficients between the concentrations of SARS-CoV-2 RNA obtained by the three assays and between the number of hospitalized COVID-19 patients and the concentration of SARS-CoV-2 RNA at each hospital.

## 3. Results and Discussion

### 3.1. Performance of Charité assays on SARS-CoV-2 quantification in wastewater

The first RT-_q_PCR assays for the detection of SARS-CoV-2 were designed at the beginning of the pandemic following the disclosure of the first SARS-CoV-2 sequence, the designated Charité assays: E_Sarbecco, RdR_p_ (P1 and P2) and N_Sarbecco (Corman *et al*., 2020). Environmental studies generally rely on the use of a single assay to determine the presence of a target (La Rosa and Muscillo, 2013). However, due to sensitivity and specificity issues the WBE studies for SARS-CoV-2 have included multiple gene targets, including the Charité (Wurtzer *et al*., 2020; Medema *et al*., 2020; Chavarria-Miró *et al*., 2020) and the CDC assays (Ahmed *et al*., 2020; Medema *et al*., 2020; Randazzo *et al*., 2020). In the 32-week study reported here, the three assays were compared with respect to detection rates and concentrations to determine the need to run all three assays in future WBE studies.

Detections of SARS-CoV-2 RNA were scarcer during the lockdown and reopening months (April-May), with discrepant results among the assays (Fig. 1A). The results of SARS-CoV-2 RNA prevalence for the three assays (*n* = 404), including below and above LOD, coincided in 193 samples. This number dropped to 80 samples when considering just samples with results above the LoD. In 116 samples, detection occurred for two assays and in 95 samples only one assay was detected.

**Fig. 1.**
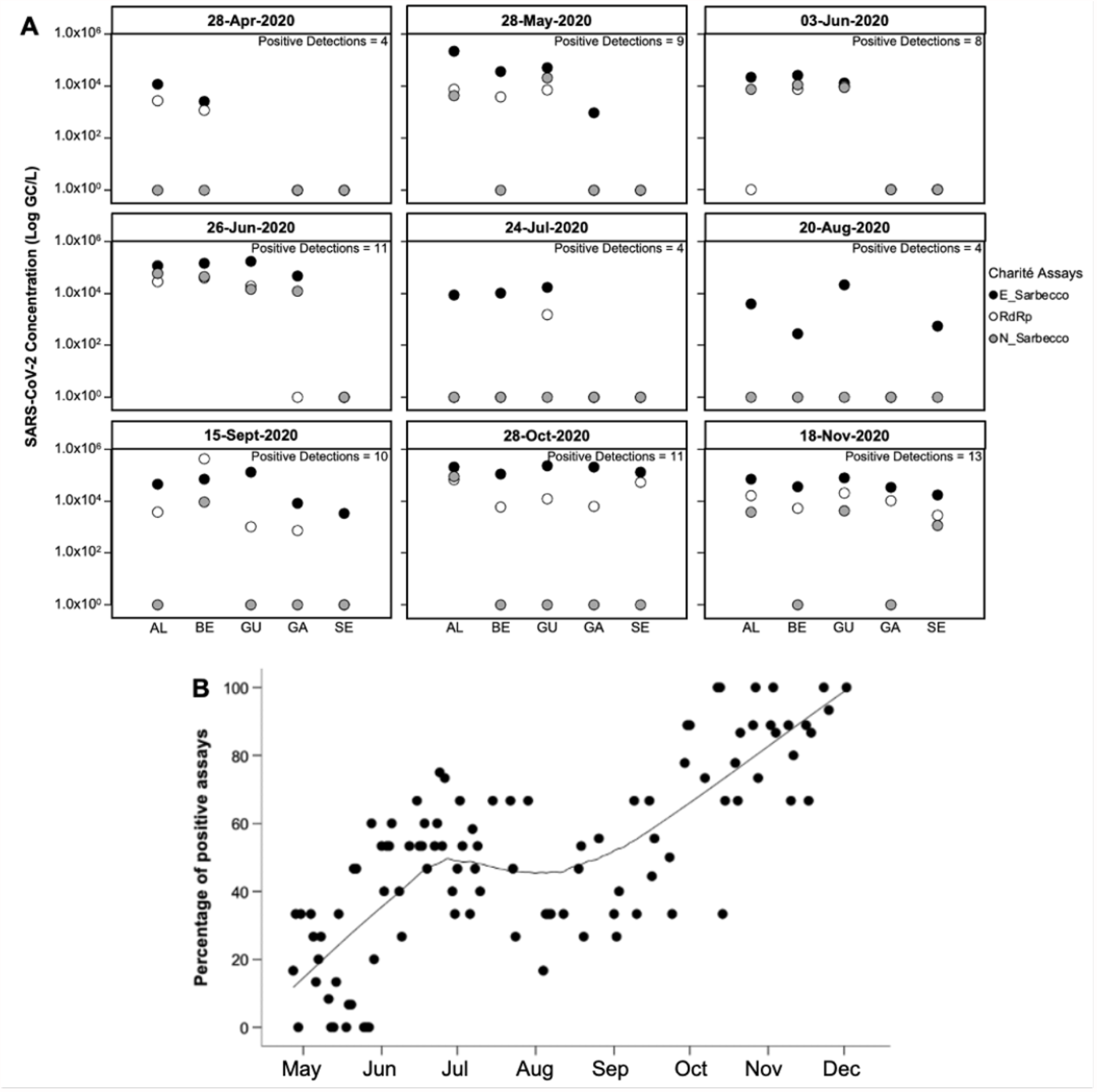
SARS-CoV-2 RNA concentration estimated with Charité assays in selected sampling dates. The concentrations in each WWTP, in selected sampling dates, are depicted on the x axis of the figure. The dates were chosen at (roughly) monthly intervals, starting from April 28, with exception of June 3, which was added because it represented one of the first dates following the complete reopening of the country (A); epidemiological phase (EPI) I: emergency state; EPI II: calamity state; EPI III: contingency and alert state; EPI IV: emergency state. Percentage of positive detection assays across the study period. Obtained with the 3 Charité assays. The trendline was drawn with LOWESS smoothing (B).

Agreement between assays increased and became more consistent as the total number of detections increased, particularly following the end of the lockdown (Fig. 1A, B). The E_Sarbecco assay was detected more frequently than the remaining assays, with consistent detections over the 32-week period of sampling. A total of 290, 177, and 100 samples tested positive for E_Sarbecco, RdR_p_, and N_Sarbecco, respectively. The detection rates for all assays showed statistically significant differences (KW = 181.45, degrees of freedom = 2, ρ<0.001). There was also statistical difference in the number of detections in the pair-wise comparison between individual assays (ρ<0.001, for all assays). The number of detections for N_Sarbecco assay was significantly lower than for the other two assays, possibly due to the higher limit of detection determined for this assay or possible loss of RNA integrity.

The positivity rates for RdR_p_ and N_Sarbecco assays increased along with increasing concentrations yielded by the E_Sarbecco assay. At concentrations between 10^2^ and 10^4^ GC/L, the positivity rate was 20% and 6% for the RdR_p_ and N_Sarbecco assays, respectively. For E_Sarbecco assay concentrations above 10^4^ GC/L, the positivity rates increased to 77% for the RdRp assay and 45% for the N_Sarbecco assay (Fig. S2).

The concentration of N_Sarbecco *versus* the other two assays in raw wastewater showed only moderate correlation (Spearman rank order correlation *r* = 0.50 for N_Sarbecco vs. RdR_p_; *r* = 0.56 for N_Sarbecco vs E_Sarbecco; ρ<0.01, *n* = 404). The correlation between E_Sarbecco and RdRp concentration was significant (*r* = 0.74, ρ<0.01, *n* = 404) (Fig. S3). Such figure facilitates the comparison of the distribution of positive and negative results for each pair of assays.

The discrepancies observed amongst E_Sarbeco, RdRp and N_Sarbeco assays agreed with previous reports, not only using the Charité assays but also the CDC protocol (Chavarria-Miró *et al*., 2020; Corman *et al*., 2020; Medema *et al*., 2020; Randazzo *et al*., 2020; Westhaus *et al*., 2020).

### 3.2. Detection of SARS-CoV-2 RNA in hospital wastewater samples

A total of 204 COVID-19 hospital wastewaters have been sampled in the 32-week study period and evaluated for the presence of SARS-CoV-2 RNA. Ninety-seven samples were positive for at least one SARS-CoV-2 assay (97/204; 48%), at concentrations ranging from 10^3^ to 10^6^ GC/L (Fig. S4). The percentage of positive samples varied from 24% (HSS) to 85% (HCC). The Cq values varied between 26.36 and 38.43 for the E_Sarbecco assay, with agreement in detection for the three assays in 62 % of the samples (including samples below the LoD) and in 21 % samples considering just samples with positive detection (*n* = 98). Although highly relevant, the number of studies reporting the specific detection of this virus in hospital wastewater is very limited (J. Wang *et al*., 2020; D. Zhang *et al*., 2020; Gonçalves *et al*., 2021). Although no quantification was made, J. Wang *et al*. (2020) and Gonçalves *et al*. (2021) reported similar Ct values to those obtained in our study. Detection frequency of SARS-CoV-2 RNA in hospital wastewater increased by the end of the study, when the number of cases in Portugal increased steeply and a high number of hospital beds were being occupied with COVID-19 patients (Fig. 2). From the end of the lockdown to schools reopening and return to partial face-to-face work (April through mid-September), the number of hospitalized COVID-19 cases decrease from an average of 60 to 3 in HSS and from 73 to 5 in HSO, increasing to 115 and 162 in November, respectively. As for HCC, the average monthly number of hospitalized COVID-19 cases remained stable from April to July (average ranging between 48 and 61 in April and June, respectively), decreasing during the month of August (30) only to increase again in September. By the end of the sampling period, the average number of hospitalized COVID-19 cases increased to 114.

**Fig. 2.**
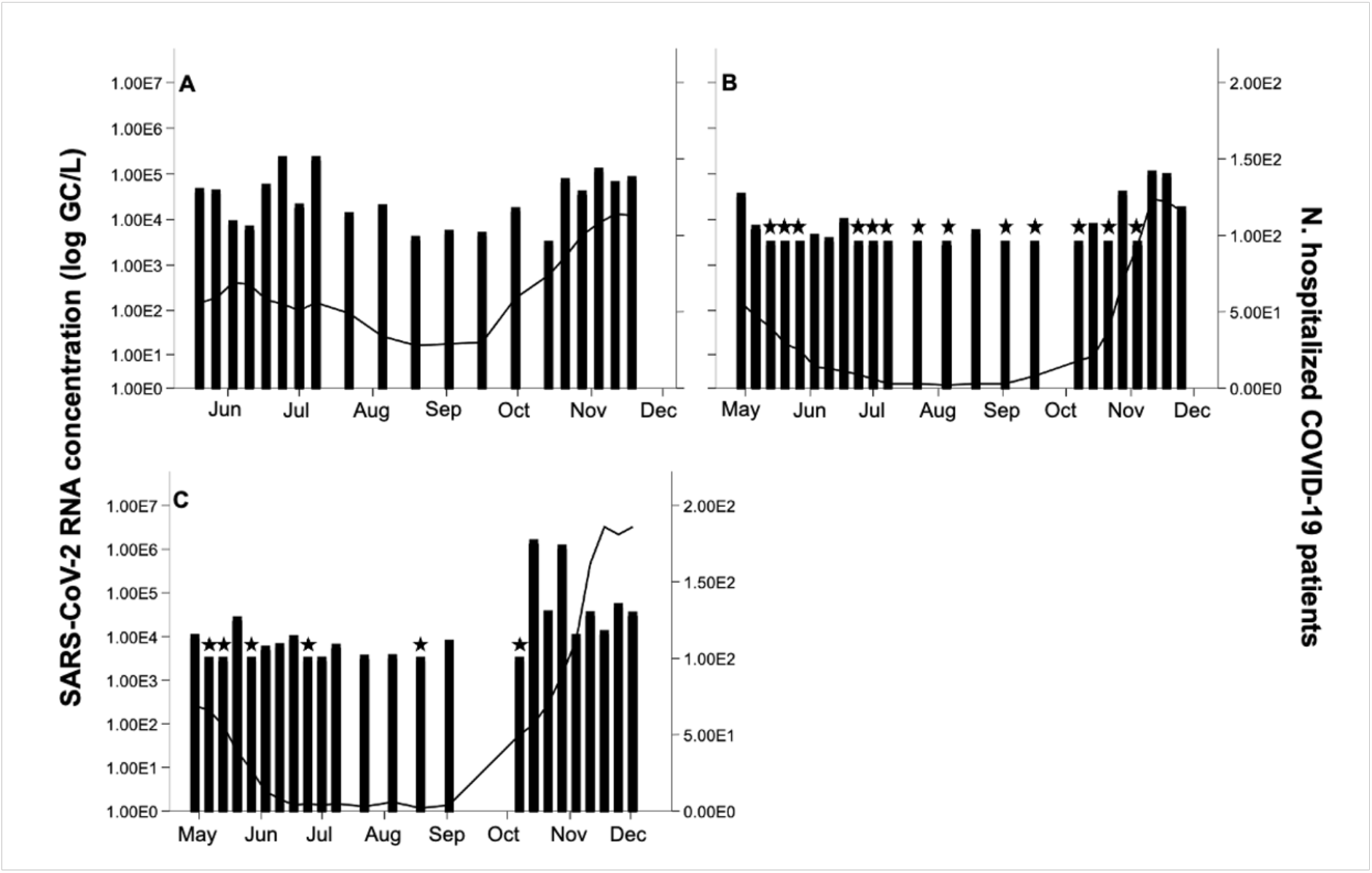
Gene fragment concentration in hospital wastewater (bars), and the number of hospitalized COVID-19 cases (line) in the three hospitals. HCC (A); HSS (B); HSO (C). ★ Indicates values below the LoD for E_Sarbecco assay. Values represented in the figures

Correlation analysis was used to investigate the quantitative relation of the SARS-CoV-2 RNA concentration to the number of hospitalized COVID-19 cases in each hospital. No correlation was found in HCC and only moderate association was obtained for the other two hospitals (Spearman rank order correlation *r* = 0.57 for HSS and *r* = 0.60 for HSO; all ρ < 0.01). During the phase with lower number of hospitalized COVID-19 cases at HSS, most of the samples collected were below the LOD, a similar result to that observed in HSO hospital (Fig. 2). On the other hand, SARS-CoV-2 RNA detection at HCC was consistent throughout the study. Sporadic detection of SARS-CoV-2 RNA during this phase could be attributed not only to the low number of hospitalized COVID-19 patients but also to the different sampling strategy. While HCC samples were composite, grab samples were taken at the other two hospitals. Statistically significant differences (ρ<0.001; Mann-Whitney U test) were determined between composite and grab samples. Composite sampling provides a better representation of a heterogenous sample than grab samples tested separately as the variance between samples decreases and the analytical results reflect more thoroughly the real composition of the sample. Automated systems (composite sampling) are commonly used for chemical analysis of water in industrial and public health applications (U.S. Geological Survey, 2006, 2010; Baird *et al*., 2017). Composite sampling has also been widely used to analyze trace contaminants such as mycotoxins in foods and to determine microbial populations in soil and water (Jarvis, 2007; Cornman *et al*., 2018). However, for quantification purposes, composite sampling has not been routinely applied in microbiological analysis of water due to a possible dilution effect. This paradigm has shifted with SARS-CoV-2, with this respiratory virus being found only in approximately 50% of the stools of infected patients at varying concentrations (10^2^ to 10^8^ per gram of stool) (Lescure *et al*., 2020; Pan *et al*., 2020; Wölfel *et al*., 2020; Y. Wu *et al*., 2020; Xu *et al*., 2020). Even if composite sampling is not paramount in WWTP settings, in single, point locations (such as hospital wastewaters) it may have a deeper impact with the results from this study corroborating the initial hypothesis, as a lower percentage of positive samples were obtained for the hospitals where grab samples were taken.

### 3.3. Temporal dynamics of SARS-CoV-2 RNA in raw wastewater

A total of 404 raw wastewater were collected between April 27 and December 2, 2020 and monitored for the presence of SARS-CoV-2 RNA. Concentration in positive samples, for E_Sarbecco assay, varied generally between 10^3^ and 10^5^ GC/L (Fig. 3).

**Fig. 3.**
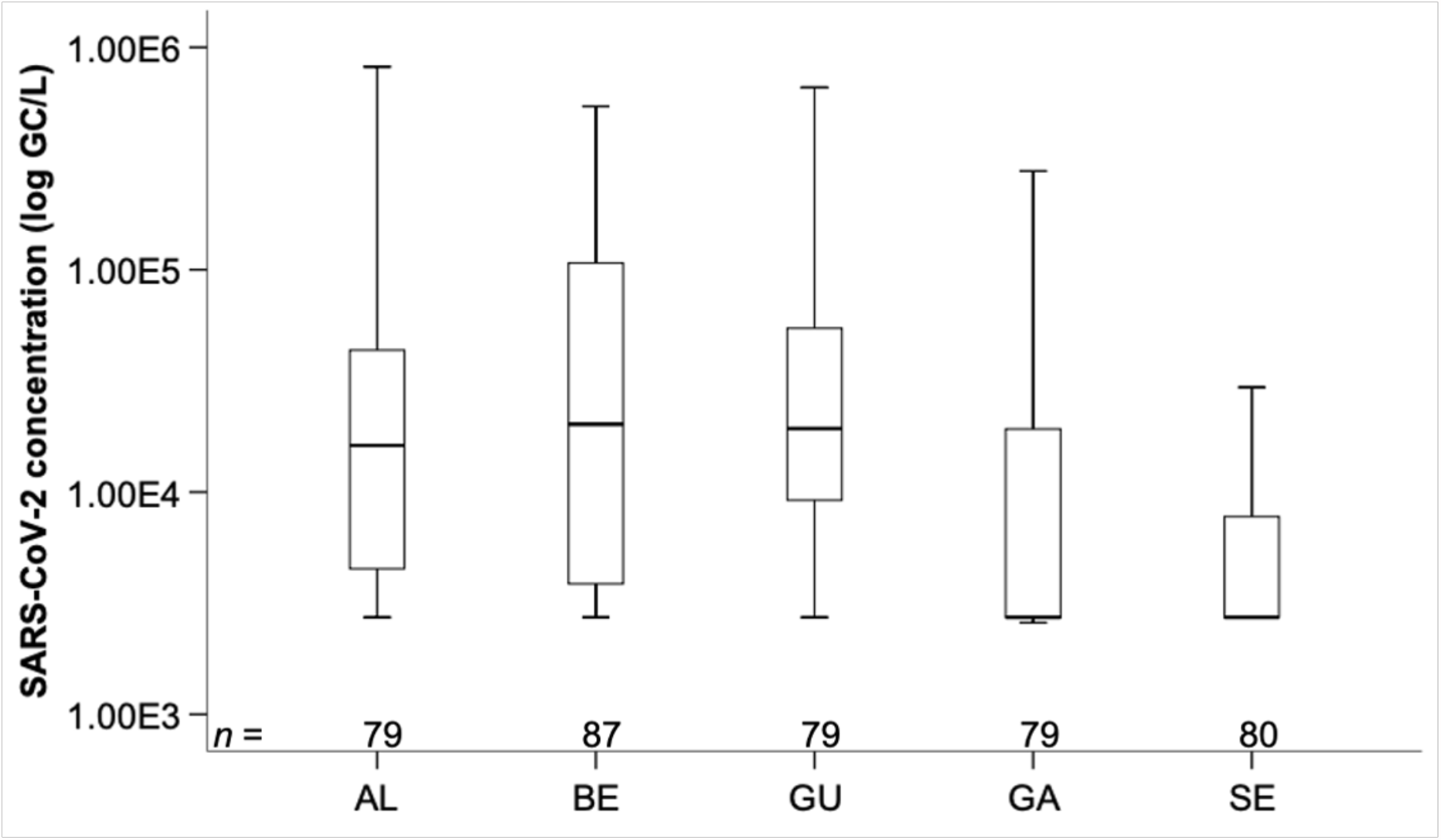
SARS-CoV-2 concentration in the tested WWTP. AL-Alcântara; BE – Beirolas; GU – Guia; GA – Gaia Litoral; SE – Serzedelo (circles and asterisks represent outliers).

Table 2 shows SARS-CoV-2 RNA concentrations and percentage of positive samples discriminated by WWTP. The prevalence of SARS-CoV-2 RNA varied between 51% in SE and 85% in BE and GU, with WWTP located in LVT conveying the highest number of positive detections.

**Table 2.** SARS-CoV-2 RNA concentration and percentage of positive samples in the overall study and in each WWTP

| Sampling location | % Positive samples | SARS-CoV-2 RNA concentration variation (GC/L) |
| --- | --- | --- |
| All WWTP | 72 (291/404) | $3.13 \times 10^3 - 8.95 \times 10^5$ |
| AL | 82 (65/79) | $3.86 \times 10^3 - 8.17 \times 10^5$ |
| BE | 85 (74/87) | $3.13 \times 10^3 - 5.43 \times 10^5$ |
| GU | 85 (67/79) | $3.41 \times 10^3 - 8.95 \times 10^5$ |
| GA | 56 (44/79) | $3.30 \times 10^3 - 3.93 \times 10^5$ |
| SE | 51 (41/80) | $3.29 \times 10^3 - 3.20 \times 10^5$ |

The concentrations found in this study are in line with those documented in the US, and The Netherlands (Gonzalez *et al*., 2020; Medema *et al*., 2020; Sherchan *et al*., 2020). Nonetheless, studies developed in Spain and France documented concentrations at least two orders of magnitude superior to the mean concentrations observed in this study (Randazzo *et al*., 2020; Wurtzer *et al*., 2020). The differences found between studies may result from a multitude of factors, including disease prevalence, but are more probably related to the variability in the workflows including detection assays.

### 3.4. Regional distribution of SARS-CoV-2 RNA concentration

This study was conducted during 32-weeks (eight months) comprising the end of lockdown (April) and consecutive reopening stages (May), full reopening with online classes for students and partial face-to-face work (June), the vacation period (July and August), schools reopening and return to partial face-to-face work (mid-September) (Fig. S5). The new number of reported cases decreased sharply from April (mean, 570) to May (mean, 249), increasing again in June (mean, 325), according to Reports from the Portuguese Health Authority (DGS, 2020). The average number of new cases decreased in July (mean, 286) and August (mean, 224) only to increase again in September (mean, 605), October (mean, 2,192) and November (5,058).

Fig. 4 shows the load of SARS-CoV-2 RNA, by date, normalized to population in the service area of each WWTP. SARS-CoV-2 RNA detection in WWTP for the LVT region showed lower percentages of detection during April-May, increase in the frequency of detection in June, decrease for the months of July, August and mid-September, and a steep increase from mid-September onwards (Fig. S6). The viral load in the LVT region in this region followed a similar trend to that of the prevalence of the virus. Nonetheless, the detection of SARS-CoV-2 RNA in WWTP from LVT region remained high after the end of lockdown. SARS-CoV-2 RNA load in the north region of the country (GA and SE) remained stable during the period comprising April to mid-September, sharply increasing afterwards following the trends observed in the syndromic surveillance (Fig. S6). Occasional detections were observed during the lockdown and following periods with a gradual increase in the frequency of detection until mid-September. Upon school reopening, return to partial face-to-face work, a steep increase occurred in the SARS-CoV-2 RNA load in all locations. During pre-lockdown and lockdown, the North region was the most affected by COVID-19, a pattern that shifted following the reopening with the great Lisbon area becoming the main contributor to the increase in the number of COVID-19 cases observed throughout May and June (Fig. S7). Altogether, the cumulative number of COVID-19 cases increased at a slow pace from the end of April until the beginning of October, with a noticeable increase at this stage mainly due to the new spike in cases registered in the North region. Overall, and until October 25, 2020, Lisbon and Sintra, both in LVT, had the highest number of confirmed COVID-19 cases (9,202 and 7,454, respectively), followed by Amadora, Loures, (3,722, and 4,164, respectively), also in the LVT region. In the North region, Vila Nova de Gaia had the highest number of confirmed cases.

**Fig. 4.**
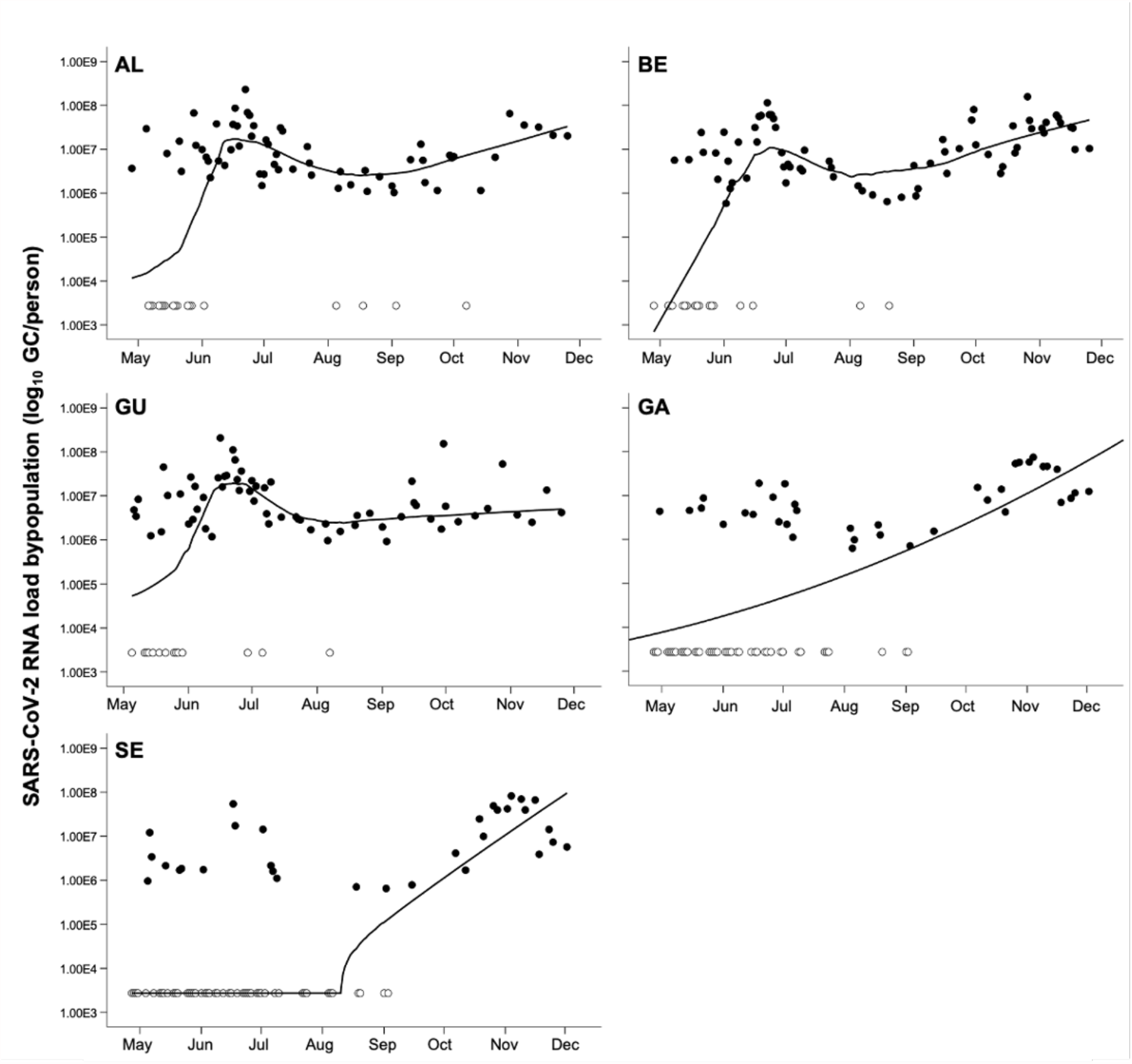
SARS-CoV-2 RNA load, by date, normalized to the population in the service area of each WWTP. Black dots indicate samples above the LoD, white dots represent samples below the LoD (with LOWESS smoothing)

Data from Fig. 4 can be used for comparison with existing outbreaks reported by the health department. For instance, the increase in the detection in the BE service area documented during June was likely caused by outbreaks in Sacavém-Prior Velho, Camarate-Unhos-Apelação and Santa Clara civil parishes. Such projection can also show trends in viruses spread over time within localized populations, not only from symptomatic but also from asymptomatic, pre-symptomatic and post-symptomatic. Such representation shows that although the number of clinically tested cases in the population was more consistent, the viral concentration remained mostly heterogeneous with a vast influence from localized hotspots of infection.

Fig. 5 illustrates the combined loads of SARS-CoV-2 RNA, over time, in the chosen WWTP service areas. The concentrations of SARS-CoV-2 RNA (E_Sarbecco) from all five WWTP were merged daily to obtain an estimation of the concentrations in the regions tested.

**Fig. 5.**
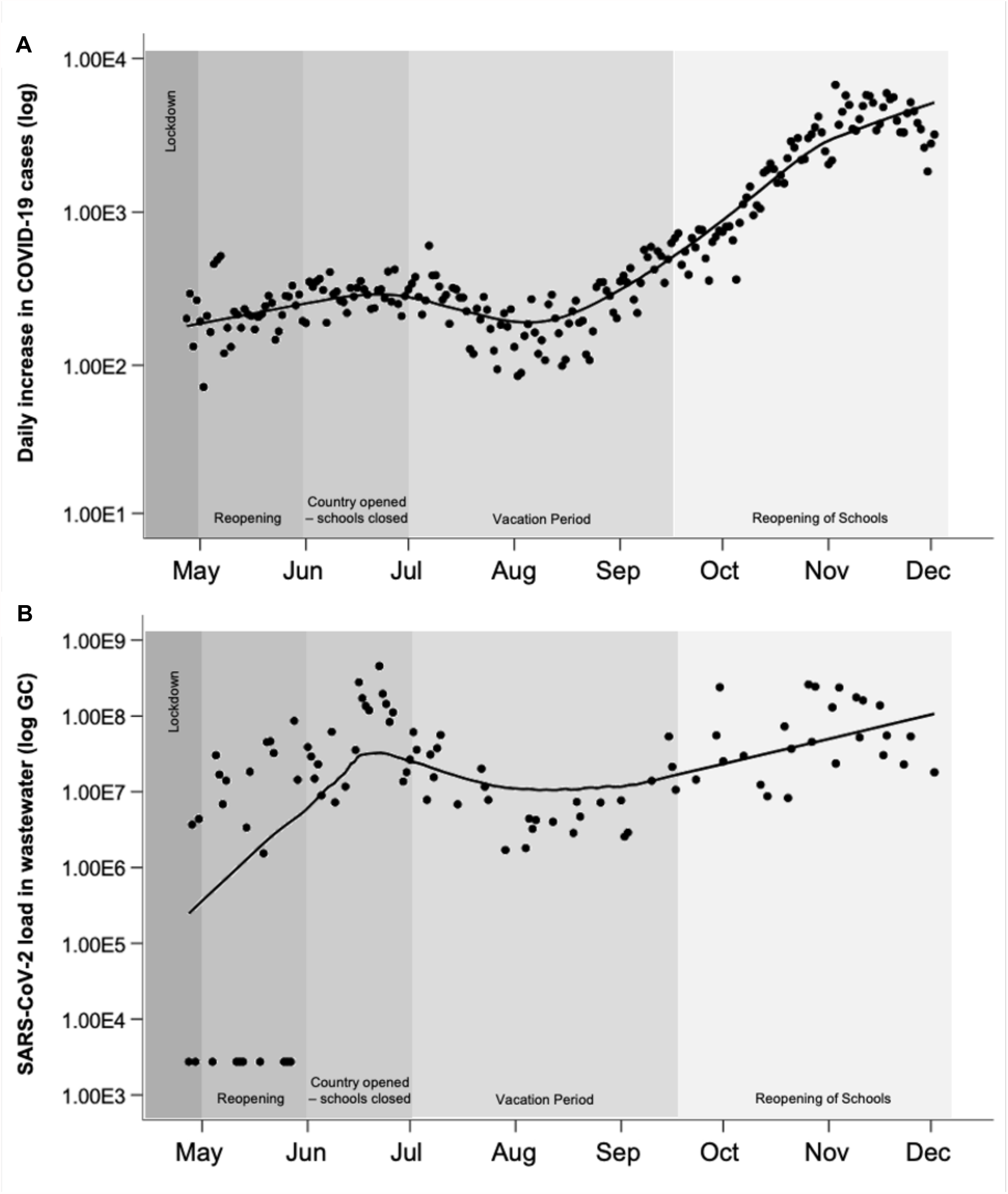
Daily increase in COVID-19 cases (A) (DGS, 2020) and combined SARS-CoV-2 concentration in wastewater for the regions under study over the 32-week period with LOWESS smoothing (B)

The trend combined for the regions was equivalent to the trends observed in the clinical surveillance. It is evident from the present data that the reopening phase, in May, corresponded to an increment in the viral load, which is in accordance with the increase observed, in Portugal, in the number of new daily COVID-19 reported cases. Following this phase, the country entered the summer vacation period, with a slight decrease on viral load. The third and final stage of viral loading, in this study, occurred after the reopening of schools and return to partial face-to-face work. At this stage, viral loading increased gradually in parallel with the rise of new daily COVID-19 cases in the country.

The pattern similarity between the number of new COVID-19 cases reported daily, provided by clinical testing, and the load of SARS-CoV-2 RNA in raw wastewater further proves the usefulness of WBE for SARS-CoV-2, or another potential future pandemic. Such representation (Fig. 5B), could therefore be integrated with syndromic surveillance data, as an early-warning system for the increase of the number of infected individuals within the community.

Results from individual testing should be the most accurate measure of transmission and disease occurrence in the population, but the scale of testing (spatial and temporal) necessary to have accurate information and to be able to follow the spread of the virus in the population is unrealistic and economically impracticable for most countries. Additionally, continuous testing indispensable for the effective control of the disease is economically and timely challenging. Wastewater monitoring represents testing thousands of infected people simultaneously rather than a single person and is complimentary to syndromic surveillance of COVID-19. The knowledge provided by the analysis of wastewater can, therefore, be employed as an impartial surveillance tool, reflecting more closely the health of a population. Moreover, wastewater may also allow for a precocious detection of new SARS-CoV-2 variants circulating in the community (Crits-Christoph *et al*., 2021; Jahn *et al*., 2021). WBE for SARS-CoV-2, and future emerging pathogens, has the potential to target the need for more localized clinical testing, facilitating the detection of occasional hotspots of infection likely to occur as this or other pandemics take place. It is scalable, with a fast turnaround, and economically competitive. WBE could be useful in school or nursing home settings, to evaluate the presence and spread of the viruses instead of testing hundreds or thousands of individuals. Additionally, WBE can be a very powerful tool in countries with limited resources, to inform decisions and in aiding with policy making

## 4. Conclusion

- SARS-CoV-2 RNA was detected in raw wastewater of all five studied WWTP at concentrations similar to those reported in other studies. Data reflected the different epidemiological stages, including surges and decreases, observed with the syndromic surveillance.
- The selection of sampling methods, composite vs grab, may have a massive impact in the results and potential use of WBE for SARS-CoV-2 or for any other future pandemic, particularly in situations where low circulation of the virus is expected.
- The total load of SARS-CoV-2 RNA in raw wastewater followed a similar trend to the number of daily new COVID-19 reported cases. Considering data, the use of viral loading would be a more suitable approach than gene-based approaches to use in WBE settings. We consider using the number of daily new COVID-19 reported cases a more suitable approach to simply comparing with cumulative number of cases especially when dealing with several waves of infection.
- Data from this study corroborates the plausibility and timeliness of the development and deployment of a nationwide WBE system for SARS-CoV-2 (naturally, ideally scalable for future pandemics) to aid local health and governmental authorities in policy making to help with future health crisis.

## Data Availability

The data contained in this manuscript has not been peer reviewed, accepted or published anywhere else.

## Funding

This work was supported by Programa Operacional de Competitividade e Internacionalização (POCI) (FEDER component), Programa Operacional Regional de Lisboa, and Programa Operacional Regional do Norte (Project COVIDETECT, ref. 048467).

## Declaration of Competing Interest

The authors declare that they have no known competing financial interests or personal relationships that could have appeared to influence the work reported in this paper.

## Acknowledgements

We thank all the workers from Águas de Portugal Group who contributed to wastewater sampling. We also thank the project’s Advisory Board (EPAL, Águas do Douro e Paiva, National Environment Agency (APA), National Health Authority (DGS) and Portuguese Water and Waste Services Regulation Authority (ERSAR).

Strategic funding of Fundação para a Ciência e a Tecnologia (FCT), Portugal, to cE3c and BioISI Research Units (UIDB/00329/2020 and UIDB/04046/2020] is also gratefully acknowledged.

## Notes

### Competing Interest Statement

The authors have declared no competing interest.

### Funding Statement

This work was supported by Programa Operacional de Competitividade e Internacionalizacao (POCI) (FEDER component), Programa Operacional Regional de Lisboa, and Programa Operacional Regional do Norte (Project COVIDETECT, ref. 048467).

### Author Declarations

All ethical guidelines have been followed and there was no need for ethics committee approval

