## Supplementary_Materials for "A Wastewater-based Epidemiology tool for COVID-19 Surveillance in Portugal"

**Table S1.** Description of the WWTPs sampled in this study.

| WWTP | Projected Flow ($m^3$/day) | Mean Flow ($m^3$/day) | Estimated Population served | Municipalities served (% of population served by WWTP) |
|---|---|---|---|---|
| Alcântara | 181,453 | 122,127 | 756,000 | Amadora (45%) |
| | | | | Lisbon (55%) |
| | | | | Oeiras (19%) |
| Beirolas | 54,500 | 36,746 | 213,510 | Lisbon (12%) |
| | | | | Loures (46%) |
| Guia | 172,800 | 185,235 | 903,000 | Amadora (46%) |
| | | | | Cascais (98%) |
| | | | | Oeiras (64%) |
| | | | | Sintra (89%) |
| Gaia Litoral | 66,700 | 36,845 | 300,000 | Vila Nova de Gaia (57%) |
| Serzedelo | 25,577 | 18,778 | 180,548 | Guimarães (50%) |
| | | | | Póvoa do Lanhoso (3%) |
| | | | | Vila Nova de Famalicão (1%) |

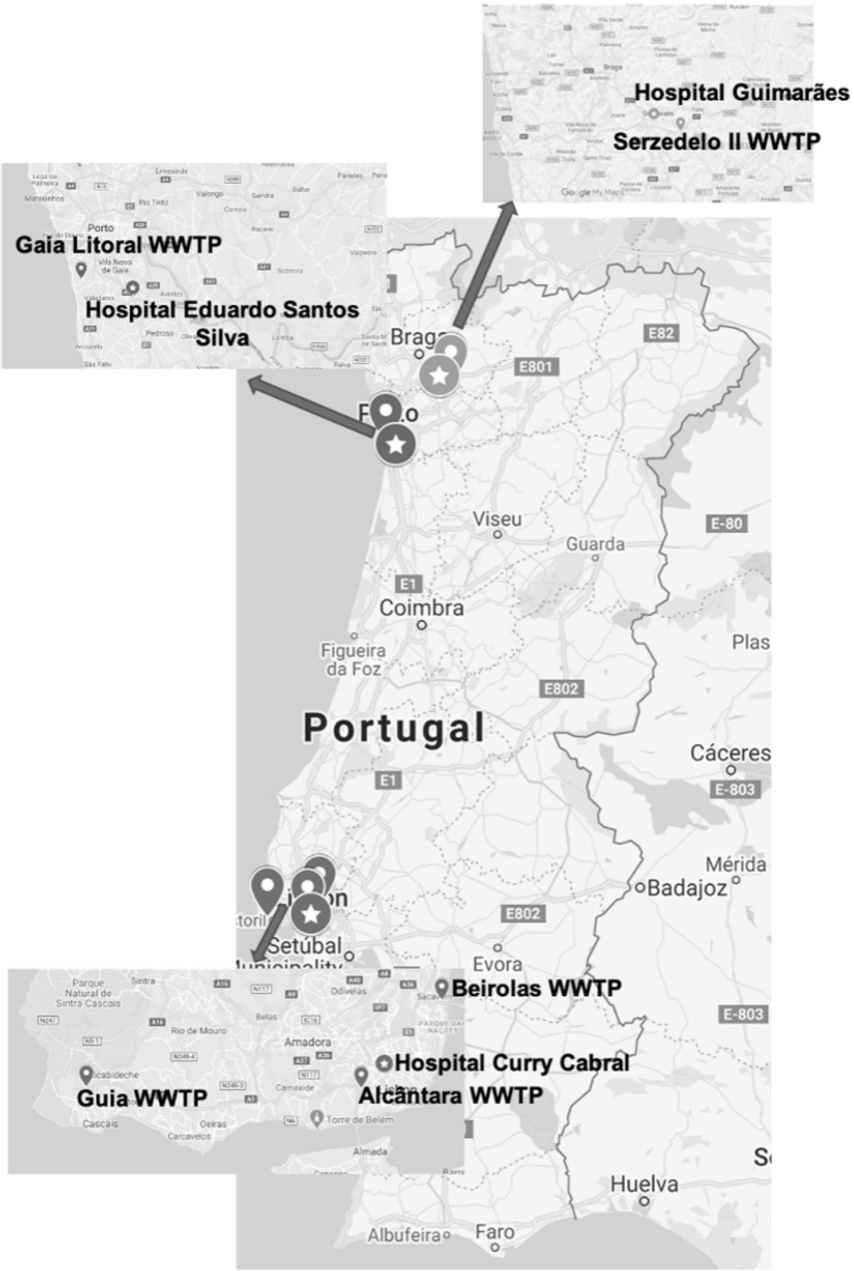


**Fig. S1.** Sampling location of the five WWTPs (bullets) and three COVID-19 hospitals (stars) monitored in this study.

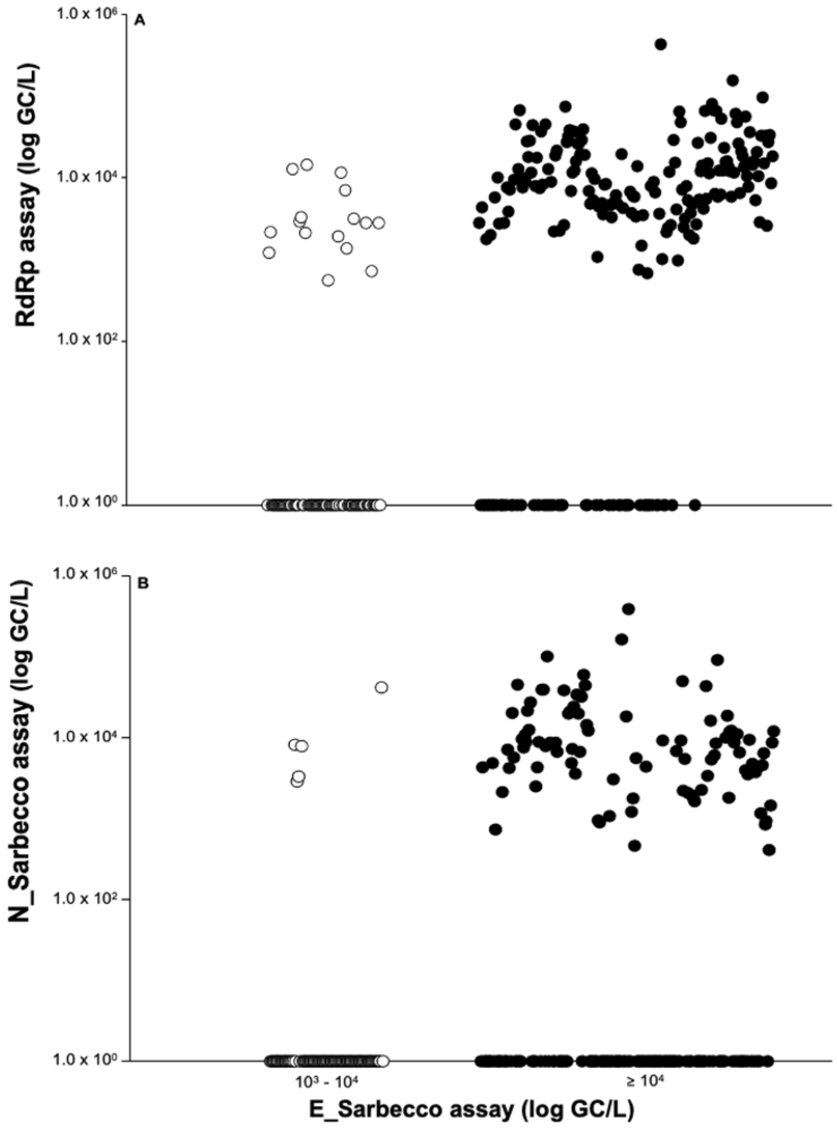


**Fig. S2.** Relationship between SARS-CoV-2 RNA concentration as detected by the E_Sarbecco assay and the levels of detection for the (A) RdRp assay and (B) N_Sarbecco assay. White dots represent samples positive for both assays, with E_Sarbecco assay concentration varying between $10^3$-$10^4$ GC/L, while black dots represent samples positive for both assays at concentrations above $10^4$ GC/L as measured by the E_Sarbecco assay.

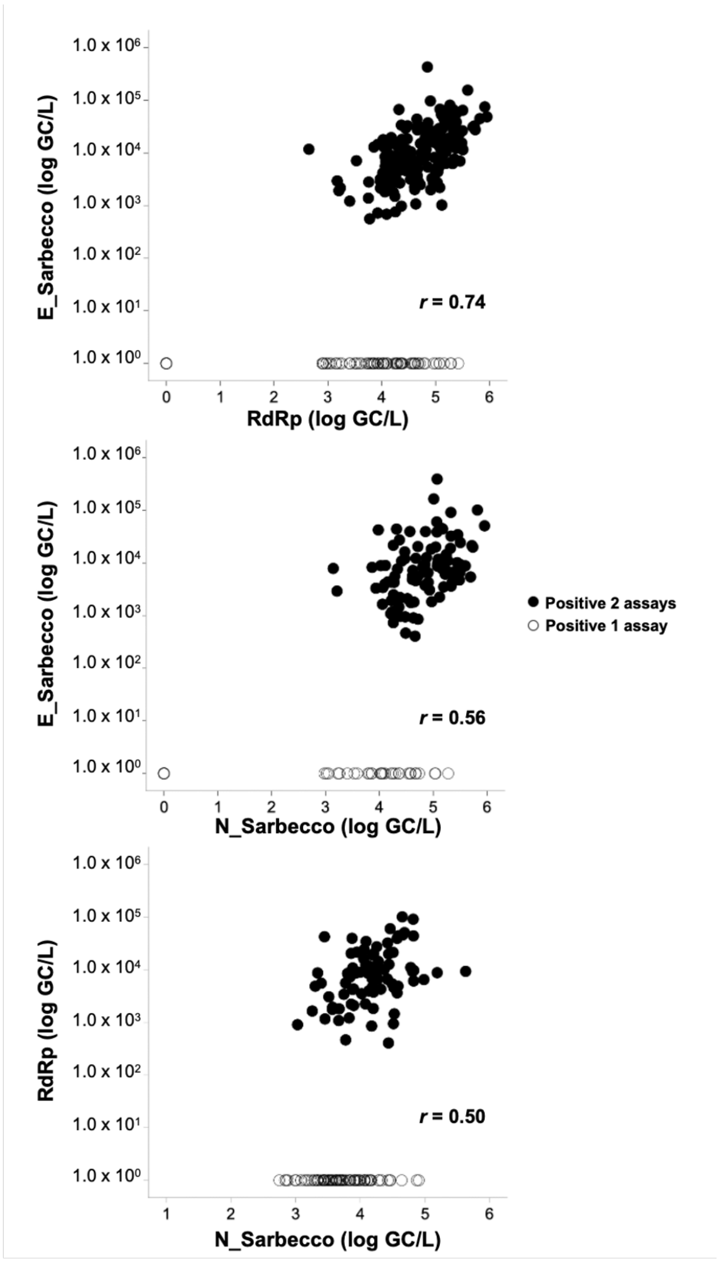


**Fig. S3.** Quantitative relationship of: E_Sarbecco assay vs RdRp assay (A), E_Sarbecco assay vs N_Sarbecco assay (B), and RdRp assay vs N_Sarbecco assay (C). Black dots correspond to samples positive for both assays, and white dots correspond to samples positive for a single assay. The coefficients for Spearman rank correlation order are given in the figure

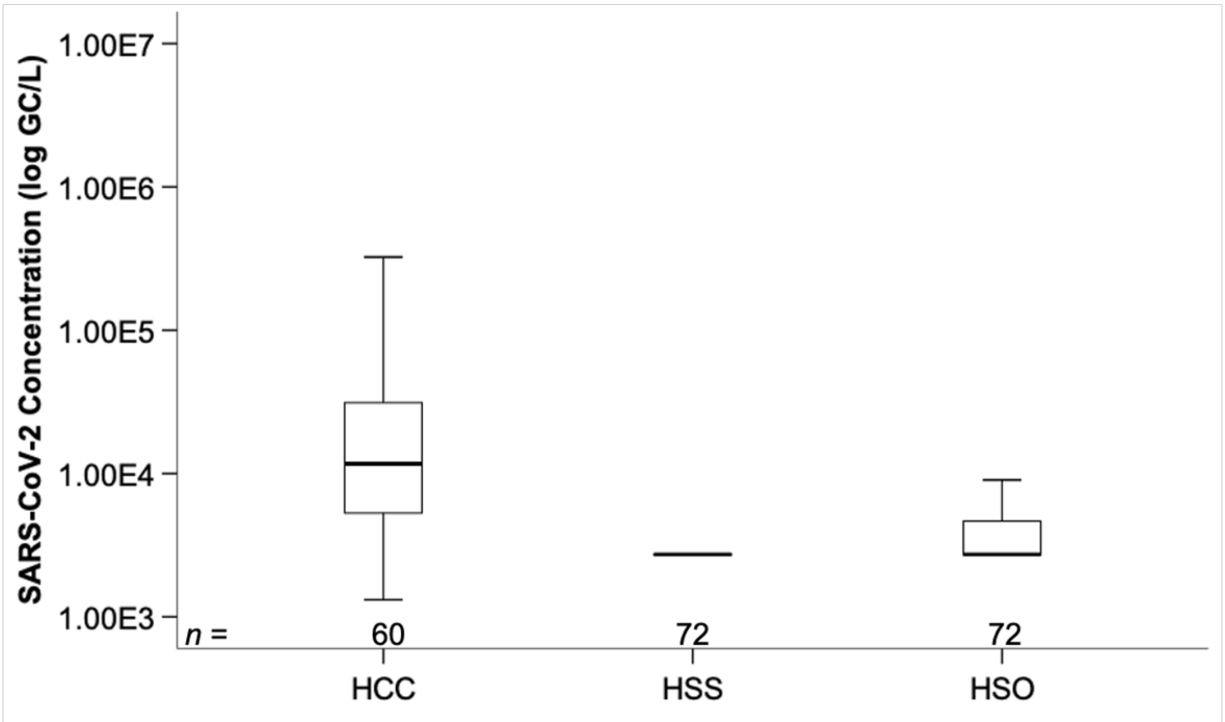


**Fig. S4.** Concentration of SARS-CoV-2 RNA determined at the three tested reference COVID-19 hospitals. HCC – Hospital Curry Cabral, HSS – Hospital Santos Silva, HSO -Hospital Sra. Oliveira.

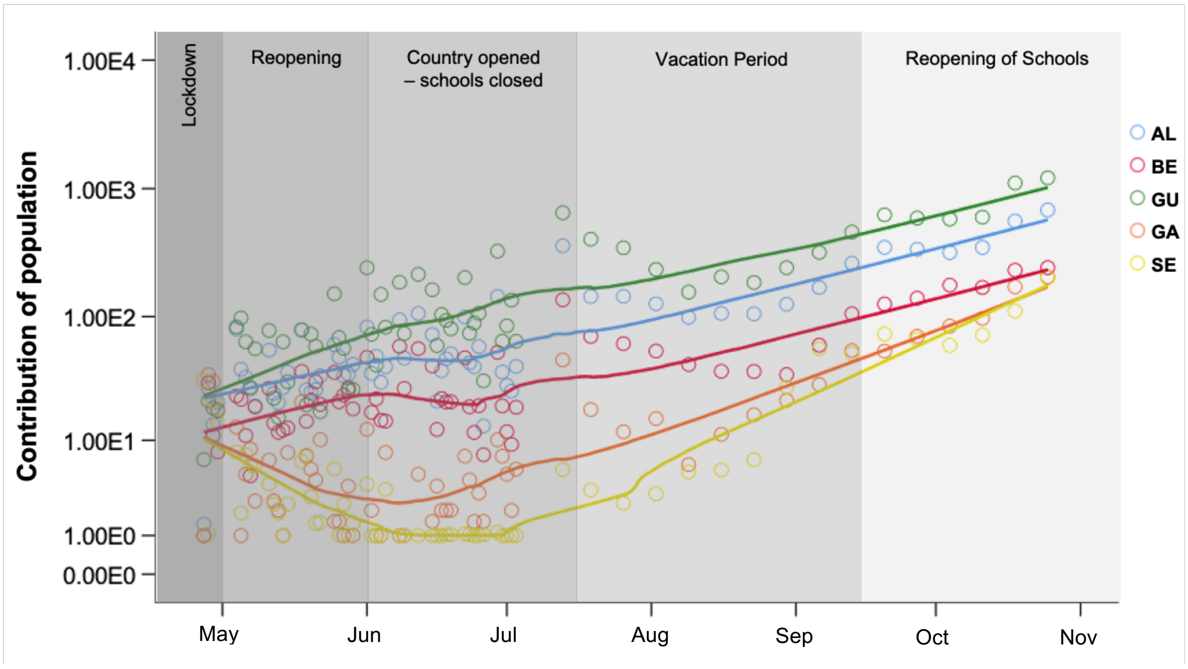


**Fig. S5.** Contribution of population to each WWTPs. The incidence rate at each WWTP was calculated by using the daily number of cases at each service area and using the percentage contribution (%) from each town to the WWTP. ALC – Alcântara; BE – Beirolas; GU – Guia; GA – Gaia Litoral; SE - Serzedelo

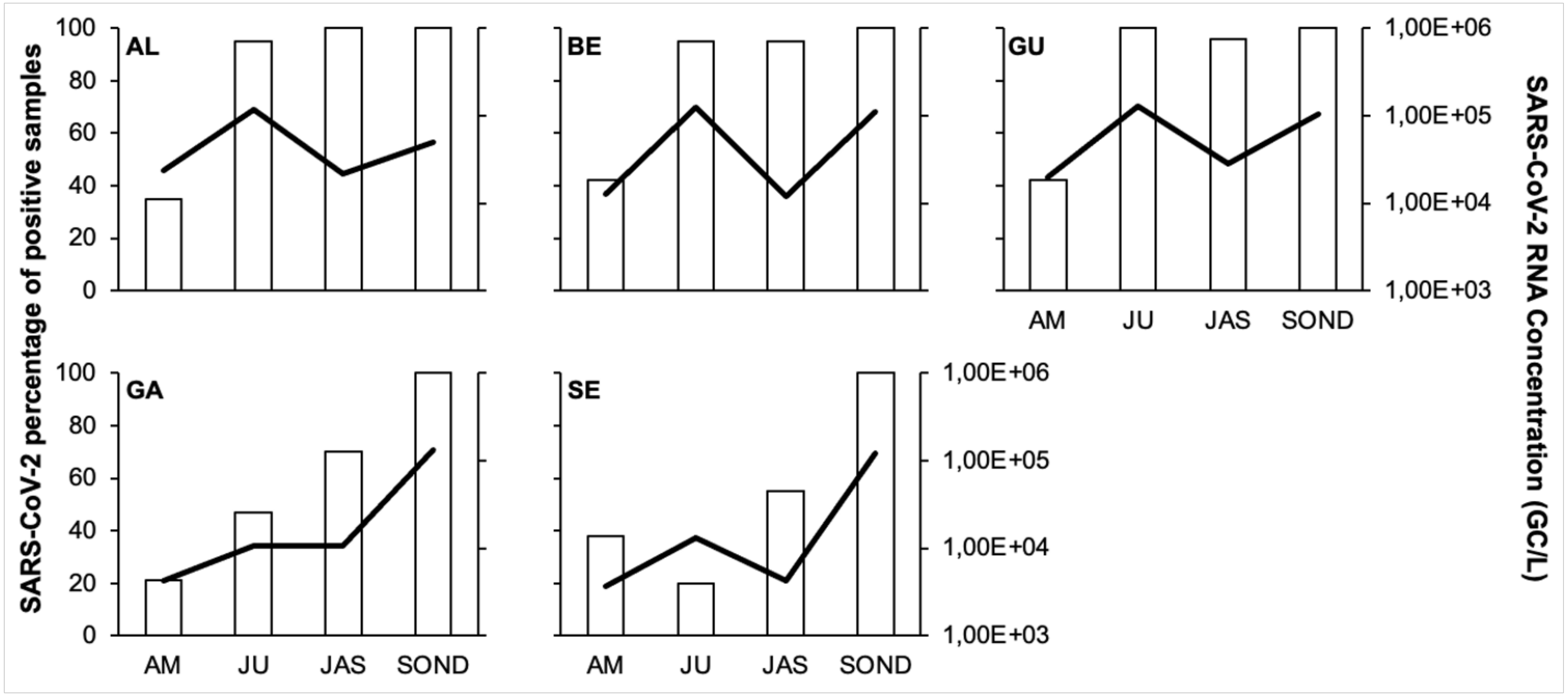


**Fig. S6.** Percentage of raw wastewater samples positive for SARS-CoV-2 and SARS-CoV-2 RNA geometric mean concentration in each WWTP (non-detected values and positive samples below the LoD were substituted by the LoD). AM- April-May; JU – June; JAS – July-August- until mid-September; SON – mid-September until the end of the study.

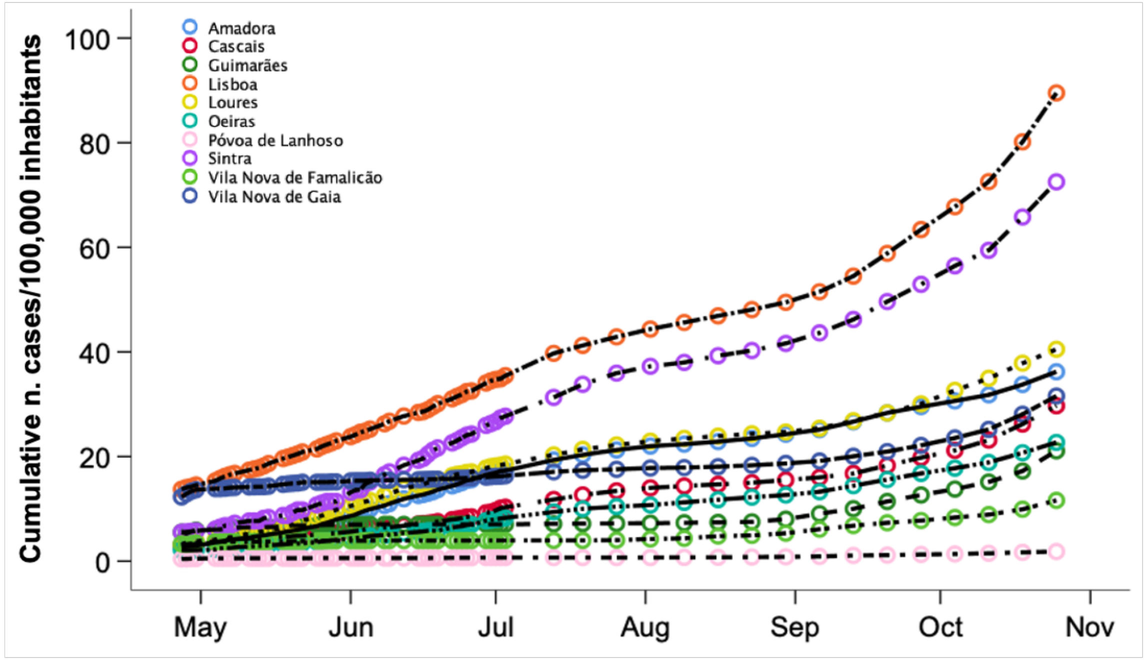


**Fig. S7.** Cumulative number of COVID-19 cases standardized by 100,000 population for the study period.
